# Glucagon-like peptide-1 receptor agonists in diabetic patients with heart failure with reduced ejection fraction

**DOI:** 10.64898/2026.11.03.26367091

**Authors:** Y. Hammer, Y. Jordi Cohen, R. Arbel, T. Razi, D. Netzer, T. Ben Gal, J.M. Weinstein, Z. Iakobishvili, D. Yamin

## Abstract

**Aims:** To evaluate the effectiveness of GLP-1 RAs on clinical outcomes in diabetic patients with heart failure with reduced ejection fraction (HFrEF).

**Methods and results:** In this retrospective observational cohort study, all adults with diabetes mellitus and HFrEF between 2014 and 2025 were identified. Patients treated with GLP-1 RAs were matched to controls using stratified exact matching. The primary outcome was a composite of heart failure hospitalization or all-cause mortality. Treatment effects were estimated using nonparametric bootstrap resampling (1,000 iterations) to derive risk ratios, NNT estimates, and corresponding 95% confidence intervals. Time-to-event analyses using Cox proportional hazards models were performed as a validation analysis. Among 7,878 patients with HFrEF, 793 (10.1%) patients who initiated GLP-1 RA therapy after HF diagnosis were matched to 1,825 controls. GLP-1 receptor agonist use was associated with a lower risk of heart failure hospitalization or death, with risk ratios of 0.58 (95% CI: 0.42–0.77) at 12 months and 0.72 (95% CI: 0.61–0.85) at 36 months. The combination of high baseline event rates and substantial relative risk reduction translated into remarkably low NNT values in the overall population, with an NNT of 17 (95% CI: 11–33) at 1 year and 13 (95% CI: 8–25) at 3 years. Post hoc subgroup analyses demonstrated consistent effects across clinical subgroups, with no significant heterogeneity observed.

**Conclusion:** In this large real-world cohort of diabetic patients with HFrEF, GLP-1 receptor agonist therapy, in addition to guideline-directed medical therapy, was associated with a lower risk of death or heart failure hospitalization.

**Graphical abstract:** 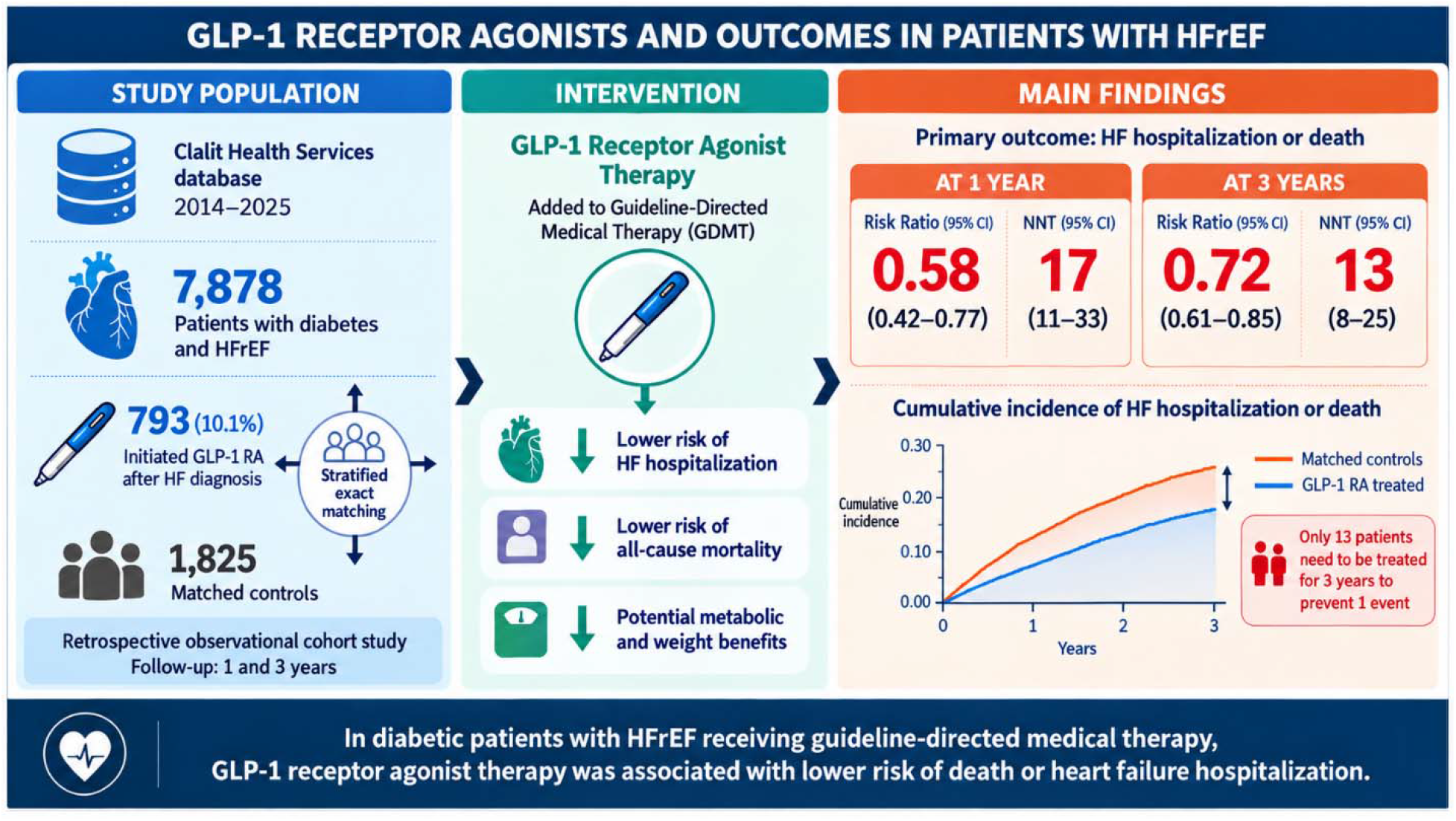

## Introduction

Despite substantial advances in pharmacological therapies for heart failure with reduced ejection fraction (HFrEF) over the past two decades, patients with this condition continue to experience high rates of mortality and heart failure hospitalization, along with impaired quality of life [1].

Glucagon-like peptide-1 receptor agonists (GLP-1 RA) include several medications which bind to a GLP-1 receptor and thereby promote insulin secretion, inhibit glucagon release and gastric emptying, and induce satiety [2]. Various GLP-1 RA differ in their structure and therefore differ in their dose, duration of action, mode of administration, and efficacy [3]. GLP-1 RA therapy has been investigated in several cohorts and was shown to decrease the likelihood of death and major adverse cardiac events as well renal outcomes [4–9]. However, as the prevalence of HF ranged from 9-24% in those studies, evidence regarding the effect of GLP-1 RA in patients with HF in general and HFrEF in particular, is scarce. While the recent publication of the STEP-HFpEF trials have provided evidence regarding the possible beneficial effect of GLP-1 RA in patients with heart failure with preserved ejection fraction (HFpEF) [10,11], there is still no robust evidence regarding the efficacy in those with HFrEF, as only 3 small clinical trials have examined the use of GLP-1 RA in patients with HFrEF [12–14], and none currently in progress.

Furthermore, while GLP-1 RA do not feature as recommended therapy in patients with HFrEF per latest published HF guidelines [15,16], a substantial number of patients with HFrEF are treated with GLP-1 RA for indications other than HF. Therefore, the aim of this study was to evaluate the real-world effectiveness of GLP-1 receptor agonist therapy on the risk of heart failure hospitalization and all-cause mortality in patients with HFrEF.

## Methods

### Study Population and Data Sources

The study was approved by the Helsinki institutional review board of Clalit Health Services. This retrospective cohort study was based on the Clalit Health Services database, the largest healthcare organization in Israel, providing universal healthcare coverage to more than 5 million members, including approximately two-thirds of the population aged >65 years. Demographic, social, clinical, medication, and laboratory data were extracted from the electronic medical records of eligible patients. Left ventricular ejection fraction (LVEF) was obtained from echocardiographic reports to identify patients with HFrEF. Eligible patients were adults aged 50 and above with diabetes mellitus, a validated ICD-9 diagnosis of heart failure (codes 428.x), and documented LVEF ≤ 40%. Patients were subsequently categorized according to GLP-1 receptor agonist exposure status. The GLP-1 receptor agonist group included patients treated with liraglutide, exenatide, dulaglutide, or semaglutide, defined by at least 3 consecutive dispensed prescriptions. To evaluate the association of GLP-1 receptor agonist therapy initiated after established HFrEF diagnosis, only patients who started GLP-1 receptor agonist treatment after the date of HFrEF diagnosis were included in the GLP-1 receptor agonist group. The comparison group consisted of patients with HFrEF who were not treated with GLP-1 receptor agonists. Exclusion criteria included prior GLP-1 receptor agonist use before HFrEF diagnosis, heart failure with preserved ejection fraction, hemodialysis, kidney transplantation, heart transplantation, or left ventricular assist device implantation.

### Outcome Measures

The primary outcome was a composite of first heart failure hospitalization or all-cause mortality, assessed at 12 and 36 months after the prespecified 3-month post-index landmark. Heart failure hospitalizations were identified from validated hospitalization records in the Clalit Health Services database, and mortality was ascertained from the national death registry linked to the electronic medical record. The individual components of the composite outcome were also analyzed separately. Treatment effects were summarized using risk ratios, relative and absolute risk reductions, and number needed to treat (NNT). Subgroup analyses evaluated the consistency of treatment effects across demographic characteristics, renal function, glycemic status, obesity, ischemic heart disease, and background guideline-directed medical therapy.

### Data analysis

Baseline characteristics were summarized descriptively and reported as median (interquartile range [IQR]) for continuous variables and percentages with 95% confidence intervals for categorical variables. We define index date as the day where patients received their first GLP-1 therapy We applied a 3-month post-index landmark, requiring both treated patients and matched controls to remain alive and free of the primary outcome, to mitigate the healthy initiator effect and other peri-initiation selection biases that may favor patients able to initiate GLP-1 therapy. This period also allowed sufficient time for dose titration and early treatment effects to emerge. Patients initiating GLP-1 receptor agonist therapy were matched to nonusers using stratified exact matching based on clinically categorized baseline covariates, including age (49–60, 60–70, 70–80, and >80 years), sex, body mass index (BMI; 18.5–25, 25–30, 30–40, and >40 kg/m²), estimated glomerular filtration rate (eGFR; <30, 30–45, 45–60, and >60 mL/min/1.73 m²), and HbA1c (<6.5%, 6.5–8%, and >8%) and tenure group in the diseases (0-2, 2-5, 5+ years) at the index date.

To estimate treatment effects while accounting for variability in the matching process, a repeated nonparametric bootstrap resampling framework was applied with 1,000 iterations. In each iteration, GLP-1 receptor agonist initiators were randomly sampled and matched to eligible controls within predefined strata, generating repeated matched cohorts. Event rates for the composite outcome of heart failure hospitalization or all-cause mortality were calculated at 12 and 36 months. For the primary composite outcome, each patient contributed a single event indicator based on the occurrence of either first heart failure hospitalization or all-cause mortality during the prespecified follow-up period. To further characterize the composite outcome, we conducted an additional supplementary analysis in which first heart failure hospitalization and all-cause mortality were evaluated separately (Supplementary Table S1 and Supplementary Figure S1). For the heart failure hospitalization analysis, only the first hospitalization was considered; death before hospitalization resulted in matched-pair censoring, whereby both the patient who died and their matched counterpart were censored at the time of death. Treatment effects were summarized as risk ratios, relative risk reduction, absolute risk reduction, and number needed to treat (NNT), with corresponding 95% confidence intervals derived from the bootstrap distribution.

To assess the robustness of the primary findings using an independent analytical framework, we conducted a validation analysis based on time-to-event methods using multivariable Cox proportional hazards regression models. To create a comparable cohort using a different matching strategy, we first performed propensity score matching with a nearest-neighbor approach based on sex, age at heart failure diagnosis, year of heart failure diagnosis, and sociodemographic characteristics. We then fitted Cox models stratified by matched pair and adjusted for age at index date, socioeconomic status, body mass index, renal function, glycemic status, ischemic heart disease, and background guideline-directed medical therapy. Hazard ratios (HRs) with 95% confidence intervals were estimated at 1 and 3 years of follow-up.

Post hoc subgroup analyses were performed according to sex, age, BMI, ischemic heart disease, eGFR and background medical therapies including ARNI, beta-blockers, MRA, SGLT2 inhibitors, metformin, insulin, DPP4 inhibitors. Statistical analyses were performed using Python version 3.9.7 (Python Software Foundation).

Post hoc subgroup analyses were performed to evaluate the consistency of associations between GLP-1 receptor agonist therapy and the primary outcome across clinically relevant patient subgroups. Subgroups were defined according to sex, age, body mass index (BMI), eGFR, glycemic status, ischemic heart disease, and background medical therapies, including ARNI, Insulin, MRA, Metformin, SGLT2i, beta-blockers, mineralocorticoid receptor antagonists (MRA), SGLT2 inhibitors, insulin, metformin, and DPP-4 inhibitors. Treatment effects within subgroups were summarized using risk ratios with corresponding 95% confidence intervals at 12- and 36-month follow-up periods. Relative risk reduction, absolute risk reduction, and number needed to treat (NNT) were additionally calculated for subgroup analyses.

## Results

### Patient population

Out of 7,878 individuals identified in the initial cohort with DM and HFrEF, 821 (10.4%) were excluded because of dialysis, prior kidney transplantation, or an estimated glomerular filtration rate (eGFR) <15 mL/min/1.73 m² (figure 1). Among the remaining 7,057 eligible individuals, 1,519 (21.5%) were excluded because of GLP-1 receptor agonist use before HFrEF diagnosis. The final study cohort therefore included 5,538 patients.

**Figure 1.**
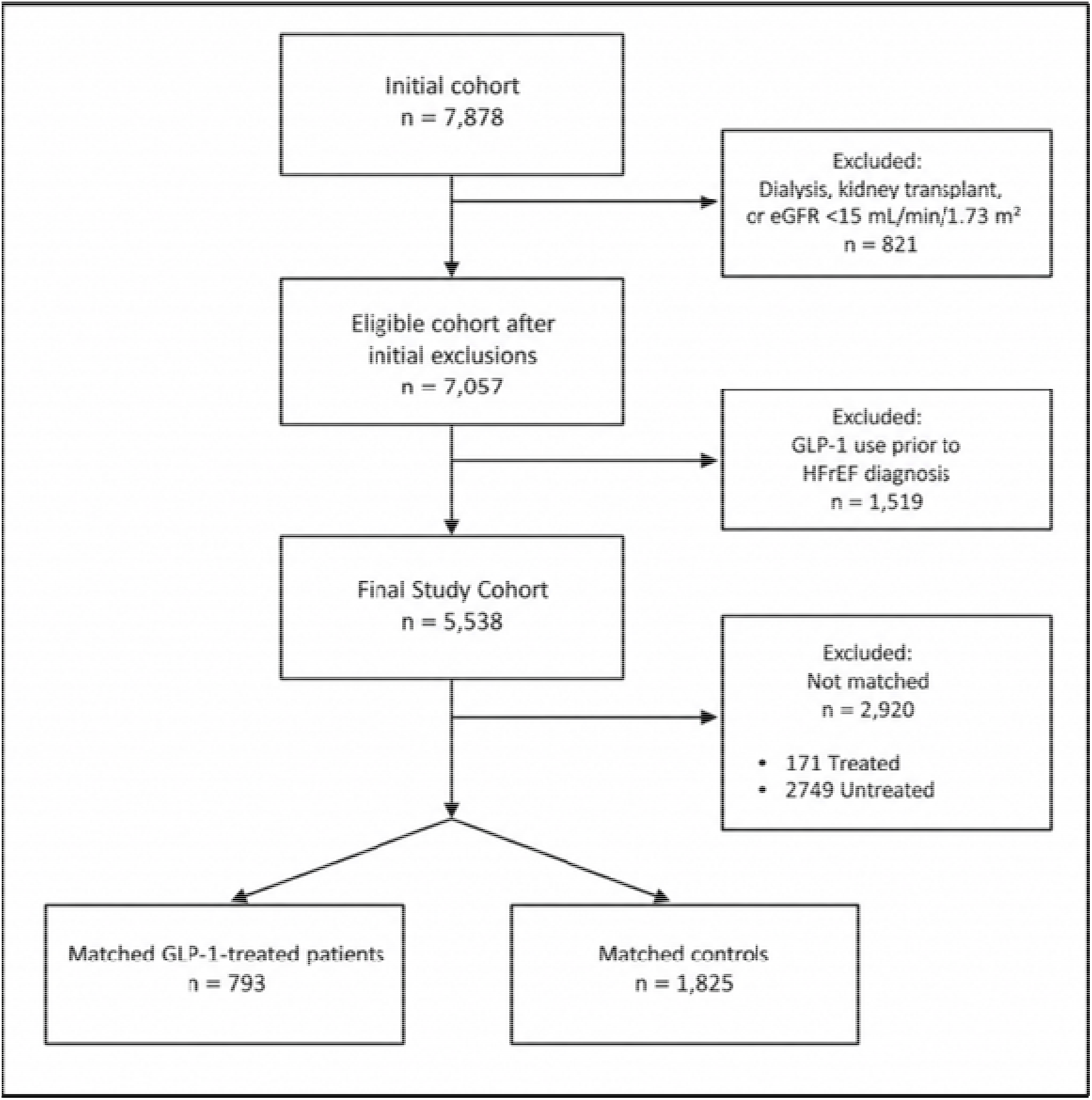
Study cohort selection and matching process for patients with HFrEF treated with GLP-1 receptor agonists and matched controls.

We successfully matched 793 of 964 (82.3%) GLP-1 receptor agonist initiators to 1,825 controls. Baseline characteristics were broadly similar between GLP-1 receptor agonist initiators and matched controls (Table 1). Median age was comparable between groups (65.0 [IQR: 60.0–70.0] years vs 67.0 [IQR: 62.0–73.0] years), with most patients aged 60–70 years, and the proportion of male patients was similar (79.7% vs 80.3%). Renal function distribution was also comparable, with most patients having an eGFR ≥60 mL/min/1.73 m². Use of guideline-directed medical therapy, including ACEi/ARB and beta-blockers, was high in both groups. GLP-1 receptor agonist initiators had numerically higher use of several cardiometabolic therapies, including SGLT2 inhibitors, mineralocorticoid receptor antagonists, insulin, and DPP-4 inhibitors. Overall, these findings suggest that GLP-1 receptor agonist initiators did not represent a clinically lower-risk population at baseline.

**Table 1.** Baseline demographic, clinical, laboratory, and treatment characteristics of GLP-1 receptor agonist initiators and matched controls. Values are presented as mean percentages with 95% confidence intervals across matched bootstrap cohorts unless otherwise indicated. Continuous variables are presented as median [interquartile range, IQR]. GLP-1 RA = glucagon-like peptide-1 receptor agonist; BMI = body mass index; eGFR = estimated glomerular filtration rate; HbA1c = glycated hemoglobin; ACEi = angiotensin-converting enzyme inhibitor; ARB = angiotensin receptor blocker; ARNI = angiotensin receptor-neprilysin inhibitor; MRA = mineralocorticoid receptor antagonist; SGLT2 = sodium-glucose cotransporter-2 inhibitor; DPP-4 = dipeptidyl peptidase-4 inhibitor; IHD =

| Characteristic | Category | Patients treated with GLP-1 receptor agonists, mean % (95% CI)<br>[N = 793] | Matched controls, mean % (95% CI)<br>[N = 1825] |
|---|---|---|---|
| Age, median [IQR] | 65.0 [60.0, 71.0] | 65.0 [60.0, 71.0] | 65.0 [60.0, 71.0] |
| Ejection fraction % |  | 34.7% [34.1-35.4] | 34.4% [33.7-35.1] |
| Age, years | 50-60 | 24.6% (21.7 - 27.7%) | 23.3% (21.1 - 25.3%) |
|  | 60-70 | 47.9% (44.5 - 51.4%) | 47.6% (45.0 - 50.1%) |
|  | 70-80 | 27.2% (24.3 - 30.4%) | 28.8% (26.6 - 31.2%) |
|  | 80+ | 0.3% (0.1 - 0.9%) | 0.3% (0.0 - 0.5%) |
| BMI | 18.5-25 | 7.6% (5.9 - 9.6%) | 9.0% (7.4 - 10.7%) |
|  | 25-30 | 38.2% (34.9 - 41.6%) | 40.2% (37.6 - 42.8%) |
|  | 30-40 | 50.4% (47.0 - 53.9%) | 47.0% (44.5 - 49.7%) |
|  | 40+ | 3.8% (2.7 - 5.3%) | 3.7% (2.8 - 4.7%) |
| eGFR | <30 | 2.1% (1.3 - 3.4%) | 2.4% (1.6 - 3.2%) |
|  | 30-45 | 13.6% (11.4 - 16.2%) | 14.4% (12.5 - 16.2%) |
|  | 45-60 | 18.5% (16.0 - 21.4%) | 19.3% (17.1 - 21.3%) |
|  | 60+ | 65.7% (62.3 - 68.9%) | 63.9% (61.2 - 66.4%) |
| HbA1c | <6.5 | 14.4% (12.1 - 17.0%) | 17.0% (15.0 - 19.0%) |
|  | 6.5-8 | 36.7% (33.4 - 40.1%) | 40.8% (38.3 - 43.4%) |
|  | 8+ | 48.9% (45.5 - 52.4%) | 42.2% (39.7 - 44.5%) |
| Sex | Male | 79.7% (76.8 - 82.4%) | 80.3% (78.3 - 82.4%) |
|  | Female | 20.3% (17.6 - 23.2%) | 19.7% (17.6 - 21.7%) |
| ACEi/ARB | Yes | 62.2% (58.8 - 65.5%) | 61.0% (58.8 - 63.3%) |
|  | No | 37.8% (34.5 - 41.2%) | 39.0% (36.7 - 41.2%) |
| ARNI | Yes | 35.8% (32.5 - 39.2%) | 33.5% (31.3 - 35.6%) |
|  | No | 64.2% (60.8 - 67.5%) | 66.5% (64.4 - 68.7%) |
| Beta-blockers | Yes | 93.2% (91.4 - 94.9%) | 87.9% (86.5 - 89.4%) |
|  | No | 6.8% (5.1 - 8.6%) | 12.1% (10.6 - 13.5%) |
| MRA | Yes | 54.0% (50.5 - 57.4%) | 47.6% (45.3 - 49.9%) |
|  | No | 46.0% (42.6 - 49.5%) | 52.4% (50.1 - 54.7%) |
| SGLT2 inhibitors | Yes | 64.9% (61.6 - 68.3%) | 53.6% (51.4 - 54.7%) |
|  | No | 35.1% (31.7 - 38.4%) | 46.4% (44.1 - 48.6%) |
| Metformin | Yes | 75.0% (72.0 - 78.0%) | 61.1% (58.9 - 63.3%) |
|  | No | 25.0% (22.0 - 28.0%) | 38.9% (36.7 - 41.1%) |
| Insulin | Yes | 33.2% (29.9 - 36.4%) | 21.3% (19.4 - 23.2%) |
|  | No | 66.8% (63.6 - 70.1%) | 78.7% (76.8 - 80.6%) |
| DPP4 inhibitors | Yes | 45.5% (42.1 - 49.0%) | 29.9% (26.9 - 33.0%) |
|  | No | 54.5% (51.0 - 57.9%) | 70.1% (67.0 - 73.1%) |
| IHD | Yes | 88.5% (86.1 - 90.6%) | 89.9% (88.5 - 91.3%) |
|  | No | 11.5% (9.4 - 13.9%) | 10.1% (8.7 - 11.5%) |

### Clinical outcomes

GLP-1 receptor agonist use was associated with a lower cumulative incidence of the composite outcome of heart failure hospitalization or all-cause mortality compared with matched controls (Figure 2). At 12 months, the cumulative incidence was 8.2% among GLP-1 receptor agonist initiators versus 14.2% among matched controls, corresponding to a risk ratio of 0.58 (95% CI: 0.42–0.77). At 36 months, cumulative incidence increased to 20.3% in the GLP-1 receptor agonist group and 28.1% in matched controls, corresponding to a risk ratio of 0.72 (95% CI: 0.61–0.85). These treatment effects translated into remarkably low NNT values of 17 (95% CI: 11–33) at 1 year and 13 (95% CI: 8–25) at 3 years, underscoring both the high residual risk and substantial treatment benefit in this HFrEF population (Table 2). In a validation analysis employing time-to-event framework, GLP-1 receptor agonist therapy remained independently associated with a lower risk of heart failure hospitalization or all-cause mortality at both 1 year (HR 0.51, 95% CI 0.31–0.82; p=0.005) and 3 years (HR 0.65, 95% CI 0.48–0.89; p=0.006), supporting the robustness of the primary findings (SI Appendix).

**Figure 2.**
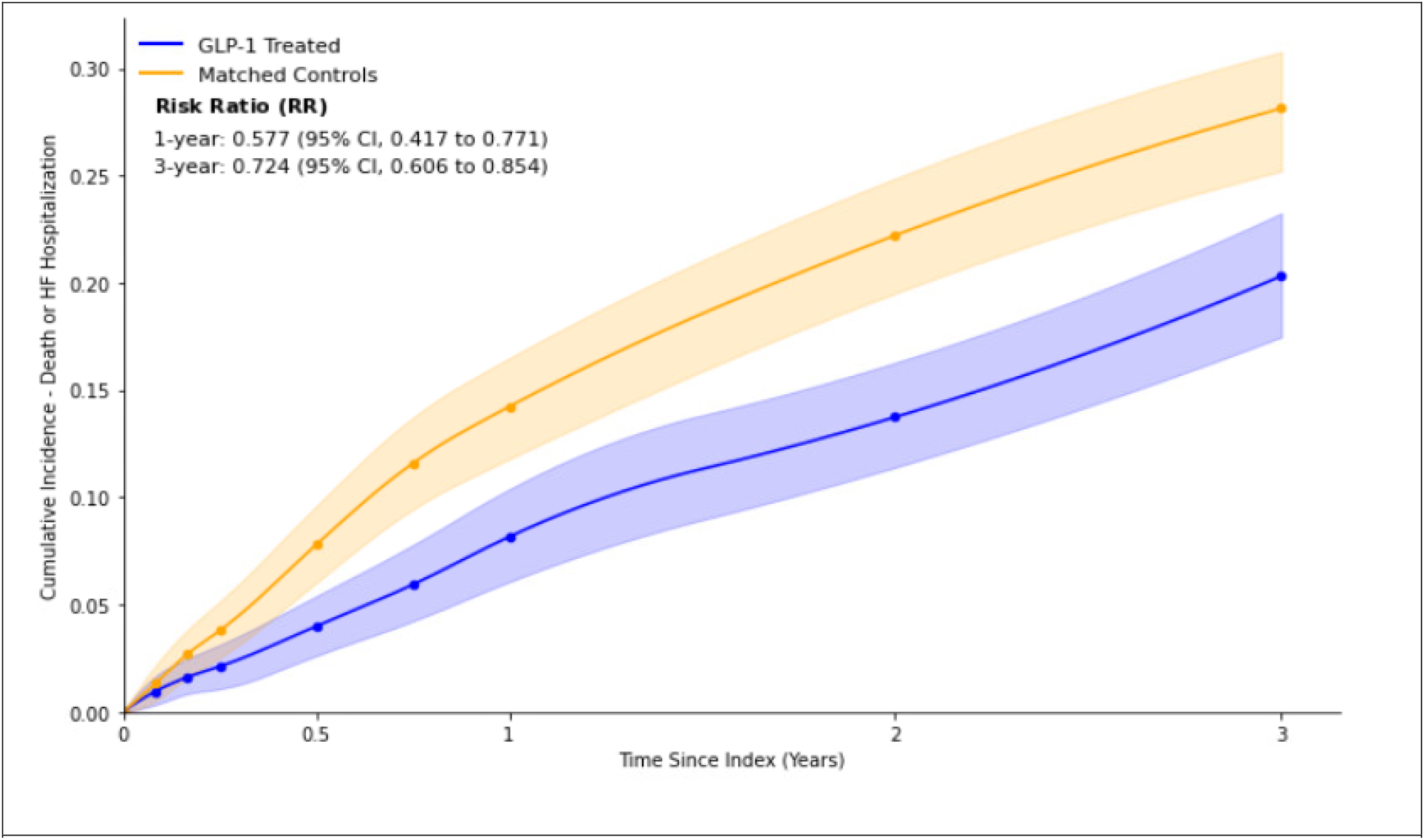
Cumulative incidence of the composite outcome of heart failure hospitalization or all-cause mortality among GLP-1 receptor agonist initiators and matched controls over 3 years of follow-up. Shaded areas represent 95% confidence intervals. Risk ratios at 1 and 3 years are shown inset.

**Table 2.** Event rates, relative and absolute risk reduction, and number needed to treat for the primary composite outcome by clinical subgroup at 1 and 3 years. Event counts represent the mean number of events per matched bootstrap cohort across 1,000 iterations and may therefore be non-integer values. Event rates, relative risk reduction (RRR), absolute risk reduction (ARR), and number needed to treat (NNT) were estimated within the repeated matching and bootstrap framework. Values in parentheses represent 95% bootstrap confidence intervals. An infinite upper confidence limit for NNT indicates that the corresponding confidence interval for ARR included zero.

| Follow-up | Subgroup | Category | GLP-1 events, mean (95% CI) | GLP-1 event rate, % (95% CI) | Control events, mean (95% CI) | Control event rate, % (95% CI) | RRR, % (95% CI) | ARR, % (95% CI) | NNT (95% CI) |
|---|---|---|---|---|---|---|---|---|---|
| 1 year | All patients |  | 47.5 (35.0 - 59.0) | 8.2 (6.0 - 10.1) | 82.3 (68.0 - 96.0) | 14.2 (11.6 - 16.3) | 42.0 (23.0 - 58.0) | 6.0 (3.1 - 9.2) | 17 (11 - 33) |
|  | Sex | Female | 5.4 (2.0 - 9.0) | 5.2 (1.9 - 8.8) | 17.8 (12.0 - 24.0) | 17.0 (11.9 - 22.2) | 69.0 (46.1 - 88.5) | 11.8 (6.1 - 17.8) | 8 (6 - 16) |
|  | Sex | Male | 41.4 (30.0 - 53.0) | 8.7 (6.2 - 11.1) | 63.5 (51.0 - 76.0) | 13.4 (10.8 - 15.9) | 34.2 (11.5 - 54.1) | 4.7 (1.4 - 8.1) | 21 (12 - 70) |
|  | BMI | <30 | 26.2 (17.0 - 36.0) | 8.8 (5.9 - 11.9) | 38.4 (28.0 - 48.0) | 12.9 (9.5 - 16.3) | 30.6 (-0.1 - 57.5) | 4.1 (0.0 - 8.7) | 24 (11 - ∞) |
|  | BMI | ≥30 | 20.6 (13.0 - 28.0) | 7.3 (4.7 - 10.0) | 42.6 (33.0 - 52.0) | 15.1 (12.0 - 18.5) | 51.2 (29.4 - 70.2) | 7.8 (3.8 - 12.0) | 13 (8 - 27) |
|  | eGFR | <45 | 12.9 (7.0 - 19.0) | 13.9 (8.1 - 20.2) | 19.4 (12.0 - 27.0) | 21.0 (14.5 - 27.8) | 31.7 (-13.3 - 64.7) | 7.0 (-2.2 - 16.0) | 14 (6 - ∞) |
|  | eGFR | ≥45 | 33.9 (23.0 - 44.0) | 7.0 (4.8 - 9.0) | 61.9 (51.0 - 74.0) | 12.7 (10.4 - 15.1) | 44.7 (25.9 - 63.4) | 5.8 (2.8 - 9.1) | 17 (11 - 35) |
|  | HbA1c | <7 | 4.4 (1.0 - 8.0) | 3.3 (0.8 - 6.1) | 16.2 (10.0 - 23.0) | 12.1 (7.9 - 16.6) | 71.8 (40.0 - 94.1) | 8.8 (3.4 - 14.3) | 11 (7 - 30) |
|  | HbA1c | ≥7 | 29.5 (20.0 - 39.0) | 9.2 (6.4 - 12.2) | 51.9 (41.0 - 62.0) | 16.2 (13.0 - 19.3) | 42.4 (18.6 - 62.3) | 7.0 (2.5 - 11.3) | 14 (9 - 40) |
| 3 years | All patients |  | 117.6 (99.0 - 134.0) | 20.3 (17.2 - 23.2) | 162.8 (145.0 - 180.0) | 28.1 (25.2 - 30.9) | 28.0 (15.0 - 39.0) | 7.8 (4.0 - 11.8) | 13 (8 - 25) |
|  | Sex | Female | 14.7 (8.0 - 21.0) | 14.0 (8.1 - 20.0) | 31.0 (23.0 - 39.0) | 29.6 (23.3 - 35.9) | 52.3 (29.2 - 72.0) | 15.6 (7.5 - 23.3) | 6 (4 - 13) |
|  | Sex | Male | 102.3 (85.0 - 119.0) | 21.6 (18.1 - 24.9) | 131.4 (115.0 - 147.0) | 27.7 (24.5 - 30.9) | 21.9 (7.8 - 35.7) | 6.1 (1.9 - 10.4) | 16 (10 - 52) |
|  | BMI | <30 | 57.2 (44.0 - 71.0) | 19.3 (15.0 - 23.5) | 78.7 (65.0 - 93.0) | 26.5 (22.3 - 30.5) | 26.9 (6.9 - 46.0) | 7.2 (1.6 - 13.2) | 14 (8 - 61) |
|  | BMI | ≥30 | 59.7 (47.0 - 72.0) | 21.2 (17.1 - 25.4) | 82.7 (72.0 - 95.0) | 29.4 (25.7 - 33.3) | 27.5 (9.1 - 43.7) | 8.2 (2.4 - 13.9) | 12 (7 - 41) |
|  | eGFR | <45 | 28.6 (20.0 - 38.0) | 30.9 (22.4 - 39.0) | 41.1 (32.0 - 50.0) | 44.4 (36.7 - 52.1) | 29.8 (5.9 - 51.2) | 13.5 (2.3 - 24.7) | 7 (4 - 44) |
|  | eGFR | ≥45 | 88.4 (73.0 - 104.0) | 18.2 (15.2 - 21.2) | 121.4 (106.0 - 137.0) | 24.9 (21.9 - 28.0) | 26.9 (11.8 - 40.6) | 6.8 (2.7 - 10.9) | 15 (9 - 37) |
|  | HbA1c | <7 | 19.6 (12.0 - 28.0) | 14.7 (9.0 - 20.7) | 31.1 (23.0 - 40.0) | 23.2 (17.5 - 29.2) | 35.8 (0.0 - 62.1) | 8.6 (0.0 - 16.3) | 12 (6 - ∞) |
|  | HbA1c | ≥7 | 68.8 (55.0 - 81.0) | 21.5 (17.5 - 25.5) | 103.4 (91.0 - 116.0) | 32.4 (28.7 - 36.0) | 33.2 (17.2 - 47.2) | 10.8 (5.1 - 16.0) | 9 (6 - 19) |

The individual components of the primary composite outcome were also examined separately. Results for first HF hospitalization and all-cause mortality are presented in Supplementary Table S1 and Supplementary Figure S1.

### Subgroup analysis

Post hoc subgroup analyses demonstrated that the association between GLP-1 receptor agonist use and lower risk of heart failure hospitalization or all-cause mortality was generally consistent across clinically relevant subgroups at both 1 and 3 years (Figure 3A and B). Estimates were broadly similar across sex, BMI, renal function, glycemic status, age, ischemic heart disease status, and background medical therapies. Numerically greater relative risk reductions were observed among women, patients with BMI ≥30 kg/m², and patients with HbA1c ≥7%. Associations were observed irrespective of background use of ARNI, SGLT2 inhibitors, MRA, insulin, or metformin, although confidence intervals crossed unity in several smaller subgroups. Overall, these findings suggest a consistent treatment effect of GLP-1 receptor agonists across a range of clinically relevant patient subgroups.

**Figure 3.**
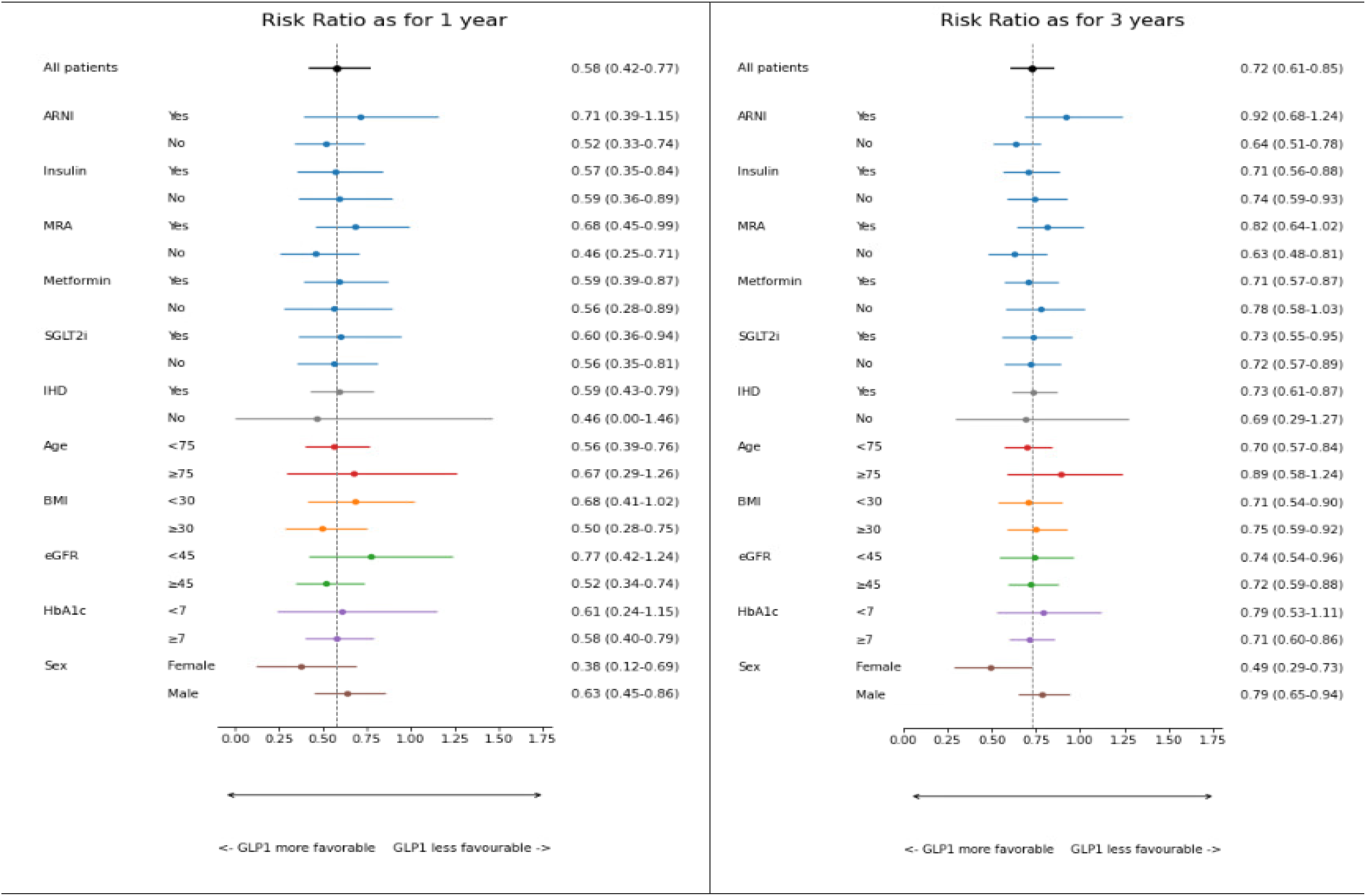
Subgroup analyses of the association between GLP-1 receptor agonist use and the composite outcome of heart failure hospitalization or all-cause mortality at 1 year (A) and 3 years (B). Forest plots display risk ratios with 95% confidence intervals across clinically relevant subgroups. The vertical dashed line represents the overall treatment effect estimate in the full study population. Values to the left of the line indicate a greater relative treatment benefit compared with the overall population estimate, whereas values to the right indicate a smaller relative treatment benefit.

Notably, several subgroups demonstrated particularly large treatment benefits or low NNT values. Women, who comprised approximately 20% of the study population, demonstrated relative risk reductions of 69.0% (95% CI: 46.1%–88.5%) at 1 year and 52.3% (95% CI: 29.2%–72.0%) at 3 years, corresponding to NNT values of 8 (95% CI: 6–16) and 6 (95% CI: 4–13), respectively (Table 2). Patients with impaired renal function (eGFR <45 mL/min/1.73 m²) demonstrated a relative risk reduction of 29.8% (95% CI: 5.9%–51.2%) at 3 years, with an NNT of 7 (95% CI: 4–44).

## Discussion

In this large real-world cohort of patients with HFrEF receiving contemporary guideline-directed medical therapy, initiation of GLP-1 receptor agonist therapy was associated with a lower risk of heart failure hospitalization or all-cause mortality at both 1 and 3 years of follow-up. GLP-1 receptor agonist use was associated with relative risk reductions of 42% at 1 year and 28% at 3 years, corresponding to absolute risk reductions of 6.0% and 7.8%, respectively. These treatment effects translated into remarkably low NNT values of 17 at 1 year and 13 at 3 years, reflecting both the substantial residual risk and clinically meaningful treatment benefit observed in this HFrEF population. Treatment effects emerged early and were sustained over time, with progressive separation in cumulative event rates translating into increasing absolute risk differences during follow-up. Importantly, these associations were consistent across a broad range of clinically relevant subgroups, with no significant heterogeneity observed, supporting the robustness and generalizability of the findings.

Given the fact that heart failure (HF) and diabetes mellitus (DM) often coexist, anti-diabetic medications have been historically prescribed to patients with cardiac disease with the purpose of lowering their glucose level and prevent long-term diabetes-related complications. Recent studies have provided evidence regarding the beneficial effect of several anti-diabetic medications that improved cardiovascular outcomes in patients with DM, independent of their glucose-lowering effect. However, while medications such as sodium-glucose cotransporter-2 (SGLT2) inhibitors have been proven to be associated with favorable clinical outcomes in patients with heart failure (both HFrEF and HFpEF) [17–20], the evidence regarding the beneficial effect of GLP-1 RA in HF in general and in HFrEF in particular, remains limited [5–8]. Furthermore, in addition to not showing any clear benefit, there was uncertainty regarding the safety of some GLP-1 RA in this patient population [13].

Several studies have examined the potential benefit of GLP-1 RA therapy in patients with HF. The 2 STEP-HFpEF trials reported improved quality of life and weight loss in a population of obese patients with HFpEF with and without DM [10,11]. The recently reported pooled analysis from 4 trials which investigated the effect of GLP-1 RA further reported a reduction in HF hospitalizations in a cohort of patients with HFpEF [21]. The evidence regarding HFrEF is not as robust. A recent study reported a reduction of death but not HF hospitalization in a large cohort of patients with HF treated with GLP-1 RA [22]. However, this study included all subtypes of HF, and therefore its results cannot be applied specifically to patients with HFrEF. Villaschi et al also reported a meta-analysis of GLP-1 RA in patients with HF (HFrEF and HFpEF) and could not find a clear benefit of GLP-1 RA in those patients. Compared with the above studies, our study focuses only on patients with HFrEF, includes a larger cohort, and displays a significant benefit of GLP-1 RA in all patients included, regardless of other factors such as age and BMI. Currently there are no randomized controlled studies to report the effect of GLP-1 RA in patients with HFrEF.

Although several bio-mechanisms have been suggested to mediate the cardiometabolic effect of GLP-1 RA, the exact mechanism by which GLP-1 RA improves quality of life and outcomes in patients with HF is still largely unknown. The recent results reported in the STEP-HFpEF trials, which included only patients with HFpEF and obesity, suggested an indirect effect through weight loss, a common comorbidity in patient with HFpEF [10]. Other suggested mechanisms for the effect of GLP-1 RA on patients with HF include anti-inflammatory and hemodynamic effects [23]. In our study, the prevalence of the primary outcomes was significantly lower in all patients treated with GLP-1 RA, regardless of their BMI. This finding suggests a different mechanism rather than pure weight loss, which will need to be further explored in future studies.

### Limitations

This study has several limitations. First, as matching was limited to selected baseline characteristics, differences between the treatment and control groups persisted, particularly with respect to medical therapy. Specifically, patients treated with GLP-1 receptor agonists were more likely to receive contemporary cardiometabolic therapies, including SGLT2 inhibitors, Metformin, insulin and MRAs.. Notably, subgroup analyses demonstrated consistent treatment effects across strata of background therapies, including SGLT2 inhibitors, MRAs, and ARNI, with no evidence of significant effect modification, supporting the robustness of the findings. Nevertheless, residual confounding cannot be fully excluded. Second, given the observational and retrospective design, causal inferences cannot be established, and unmeasured confounding and selection bias may have influenced the observed associations. Third, the database lacked detailed clinical parameters, including natriuretic peptide levels, as well as other markers of heart failure severity, limiting our ability to fully adjust for disease severity and to explore potential mechanistic pathways underlying the observed effects. Fourth, given the fact that the study population included patients with HFrEF and DM, the treatment benefit of GLP-1 RA cannot be attributable only to HFrEF, and future studies would need to examine this question in non-diabetic patients with HFrEF. Fifth, as all patients in this study were treated with GLP-1 RA for the indication of DM, medication doses were likely to be lower compared with doses prescribed nowadays for weight loss, and therefore higher does of GLP-1 RA would need to be examined in the future in this population.

### Policy implications

Although current guidelines call for caution when using GLP-1 RA in patients with HFrEF, the results of our study suggest the possible beneficial effect of GLP-1 RA as an additive therapy to the contemporary GDMT given to all patients with HFrEF. The possible addition of GLP-1 RA may further improve the clinical outcomes of this patient population by reducing the rate of death and HF hospitalization. While therapy with GLP-1 RA is associated with possible gastrointestinal side effects, they are mostly mild and tend to resolve within a short period of time, thus making the risk-benefit profile tilt towards the benefit side. Considering the already established effect of GLP-1 RA in terms of weight loss, which is a common comorbidity in HFrEF [24], GLP-1 RA may be an especially appealing treatment option for patients with HFrEF, especially in those with coexistent obesity.

## Conclusions

In this cohort study of patients with diabetes mellitus and heart failure with reduced ejection fraction, treatment with GLP-1 receptor agonists was associated with a lower risk of death or heart failure hospitalization. The observed association was consistent across clinically relevant subgroups, suggesting a potentially broad benefit and a favorable safety profile. These findings support the potential role of GLP-1 receptor agonists as an adjunct to contemporary guideline-directed medical therapy in this population. However, randomized controlled trials are warranted to confirm these findings and to better quantify the magnitude of clinical effectiveness and elucidate the underlying mechanisms of benefit.

## Data Availability

The data underlying this study were obtained from Clalit Health Services and are not publicly available due to patient privacy and confidentiality restrictions. Access to these data is subject to approval by Clalit Health Services and applicable ethical and institutional requirements.

