## Supplementary material for "Glucagon-like peptide-1 receptor agonists in diabetic patients with heart failure with reduced ejection fraction": supplementary CHF - DY Oct 6.docx

*Y. Hammer^1,2^, Y. Jordi Cohen^3^, R. Arbel^4^, T. Razi^3,4^, D. Netzer^4^, T. Ben Gal^1,2^, J.M. Weinstein^4^, Z. Iakobishvili^5-8*^, Yamin D. ^3*^*

^1^ Cardiology Department, Rabin Medical Center, Petah Tikva, Israel

^2^ Gray school of medicine, Tel-Aviv university, Tel-Aviv, Israel.

^3^ School of Industrial Engineering and Intelligent Systems, Tel-Aviv university, Tel-Aviv, Israel

^4^ Clalit Health Services, Community Medical Services Division, Tel Aviv, Israel

^5^ Cardiology Department, Soroka University Medical Center, Beer Sheva, Israel

^6^ Department of Community Cardiology, Tel Aviv Jaffa District, Clalit Health Services, Tel Aviv.
 Israel.

^7^ Department of Cardiology, Assuta Ashdod University Hospital, Ashdod, Israel.

^8^Faculty of health sciences, Ben Gurion university of the Negev, Israel.

*Equal contribution

Corresponding author:

Yoav Hammer, MD

Division of Cardiovascular Medicine, Rabin medical center, Petah-Tikva, Israel.

### Supplementary Analysis: Individual Components of the Primary Composite Outcome

To further characterize the primary composite outcome, first heart failure hospitalization and all-cause mortality were analyzed separately using the same repeated matching and bootstrap framework as in the primary analysis. Outcome follow-up began after the prespecified 3-month post-index landmark period. For the heart failure hospitalization analysis, only the first hospitalization was considered. Death before heart failure hospitalization resulted in matched-pair censoring, whereby both the patient who died and their matched counterpart were censored at the time of death. All-cause mortality was analyzed separately. Risks and mean event counts at 1 and 3 years were summarized across 1,000 bootstrap iterations, with 95% confidence intervals derived from the bootstrap distributions.

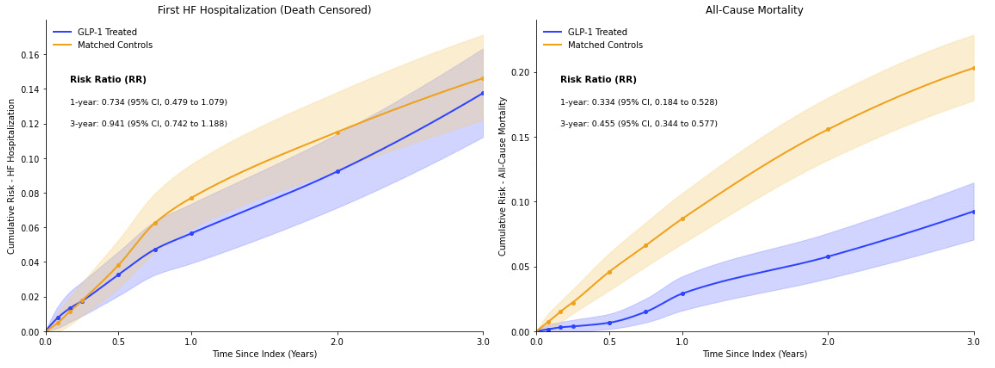

Figure S1. Individual components of the primary composite outcome. Left: cumulative risk of first heart failure hospitalization; death before hospitalization resulted in matched-pair censoring, and recurrent hospitalizations were not considered. Right: cumulative all-cause mortality. Curves show mean estimates across 1,000 matched bootstrap iterations; shaded areas indicate 95% bootstrap confidence intervals. Insets show risk ratios at 1 and 3 years.

**Supplementary Table S1. Individual components of the primary composite outcome at 1 and 3 years.**

| Outcome | Follow-up | GLP-1 mean events | GLP-1 risk, % (95% CI) | Matched controls mean events | Matched controls risk, % (95% CI) |
| --- | --- | --- | --- | --- | --- |
| First HF hospitalization, death censored | 1 year | 31.4 | 5.6 (3.9 - 7.4) | 41.4 | 7.7 (5.9 - 9.6) |
| All-cause mortality | 1 year | 16.8 | 2.9 (1.6 - 4.2) | 50.2 | 8.7 (6.7 - 10.6) |
| First HF hospitalization, death censored | 3 years | 74.7 | 13.7 (11.2 - 16.3) | 75.3 | 14.6 (12.2 - 17.1) |
| All-cause mortality | 3 years | 53.5 | 9.2 (7.1 - 11.5) | 117.6 | 20.3 (17.8 - 22.9) |

### For the first heart failure hospitalization analysis, patients were censored at the time of death if death occurred before heart failure hospitalization. Recurrent heart failure hospitalizations were not evaluated. Event counts represent the mean number of events per matched bootstrap cohort across 1,000 iterations and may therefore be non-integer values. Risks and 95% confidence intervals were derived from the corresponding bootstrap distributions.

**Supplementary Analysis: Time-to-Event Validation of the Primary Composite Outcome**

To evaluate the robustness of the primary findings using an alternative analytical framework, we conducted a validation analysis based on propensity score matching and time-to-event modelling. Propensity scores were estimated using sex, age at heart failure diagnosis (HFref), year of heart failure diagnosis, and sociodemographic characteristics. Patients receiving GLP-1 receptor agonists were matched 1:1 to untreated controls using nearest-neighbour propensity score matching with a calliper of 0.05. Following matching and data processing, the final matched cohort consisted of 878 pairs (1,756 patients).

Time-to-event analyses were performed using Cox proportional hazards regression models stratified by matched pair. In addition to GLP-1 receptor agonist treatment, models included age at index date, socioeconomic status, body mass index (<25, 25-30, 30-40, and ≥40 kg/m²), mineralocorticoid receptor antagonist (MRA) use, ACE inhibitor/angiotensin receptor blocker/angiotensin receptor-neprilysin inhibitor (ACEi/ARB/ARNI) use, SGLT2 inhibitor use, and HbA1c ≥7% as covariates.

Analyses were conducted using time-restricted Cox models at 1 year (365 days) and 3 years (1,095 days), with follow-up censored at the corresponding time horizon. GLP-1 receptor agonist therapy was associated with a lower risk of the composite outcome of heart failure hospitalization or all-cause mortality at both 1 year (HR 0.51, 95% CI 0.31-0.82; p=0.005) and 3 years (HR 0.65, 95% CI 0.48-0.89; p=0.006), demonstrating consistency with the primary analysis despite the use of a distinct matching strategy and statistical framework.

**Supplementary Table S2. Summary of Cox proportional hazards models for the primary composite outcome.**

| Follow-up | HR (GLP vs No GLP) | 95% CI | p-value | Interpretation |
| --- | --- | --- | --- | --- |
| 1 year | 0.51 | 0.31–0.82 | 0.005 | Significant reduction (~49%) |
| 3 years | 0.65 | 0.48–0.89 | 0.006 | Sustained reduction (~35%) |

**Supplementary Table S3. Multivariable Cox proportional hazards regression results in 1 and 3 years.**

| Variable | 1-year HR (95% CI) | p-value | 3-year HR (95% CI) | p-value |
| --- | --- | --- | --- | --- |
| GLP | 0.51 (0.31–0.82) | 0.005 | 0.65 (0.48–0.89) | 0.006 |
| Age | 0.94 (0.87–1.02) | 0.15 | 0.99 (0.94–1.04) | 0.6 |
| Socioeconomic status | 0.92 (0.80–1.05) | 0.2 | 0.97 (0.89–1.06) | 0.5 |
| BMI 25–30 | 0.43 (0.22–0.85) | 0.015 | 0.96 (0.62–1.48) | 0.8 |
| BMI 30–40 | 0.65 (0.31–1.36) | 0.3 | 0.97 (0.61–1.53) | 0.9 |
| BMI ≥40 | 0.23 (0.06–0.85) | 0.027 | 0.57 (0.26–1.26) | 0.2 |
| MRA | 1.2 (0.72–2.00) | 0.5 | 1.24 (0.89–1.72) | 0.2 |
| ACEi/ARB/ARNI | 1.48 (0.81–2.72) | 0.2 | 0.84 (0.55–1.27) | 0.4 |
| SGLT2 | 1.13 (0.62–2.05) | 0.7 | 0.77 (0.53–1.13) | 0.2 |
| HbA1c ≥7 | 1.42 (0.79–2.57) | 0.2 | 1.45 (0.99–2.11) | 0.057 |
